# Is COVID-19 seasonal? A time series modeling approach

**DOI:** 10.1101/2022.06.17.22276570

**Authors:** Timothy L Wiemken, Farid Khan, Jennifer L Nguyen, Luis Jodar, John M McLaughlin

**Affiliations:** Pfizer Inc. 500 Arcola Rd. Collegeville, PA 19426 USA

**Author notes:** Corresponding Author: Timothy L Wiemken Ph.D. MPH Global Clinical Epidemiologist Pfizer Vaccines, 500 Arcola Rd., Collegeville, PA 19426 USA; Alternate Corresponding Author: John M McLaughlin, PhD, Vice President, Global Franchise Lead, COVID-19 and mRNA vaccines Pfizer Inc., 500 Arcola Rd, Collegeville, PA 19426, E.

**Keywords:** SARS-CoV-2, winter, fall, autumn, surge

## Abstract

**Background:** Determining whether SARS-CoV-2 is or will be seasonal like other respiratory viruses is critical for public health planning, including informing vaccine policy regarding the optimal timing for deploying booster doses. To help answer this urgent public health question, we evaluated whether COVID-19 case rates in the United States and Europe followed a seasonal pattern using time series models.

**Methods:** We analyzed COVID-19 data from Our World in Data from Mar 2020 through Apr 2022 for the United States (and Census Region) and five European countries (Italy, France, Germany, Spain, and the United Kingdom). For each, anomalies were identified using Twitter’s decomposition method and Generalized Extreme Studentized Deviate tests. We performed sensitivity analyses to determine the impact of data source (i.e., using US Centers for Disease Control and Prevention [CDC] data instead of OWID) and whether findings were similar after adjusting for multiple covariates. Finally, we determined whether our time series models accurately predicted seasonal influenza trends using US CDC FluView data.

**Results:** Anomaly plots detected COVID-19 rates that were higher than expected between November and March each year in the United States and Europe. In the US Southern Census Region, in addition to seasonal peaks in the fall/winter, a second peak in Aug/Sep 2021 was identified as anomalous. Results were robust to sensitivity analyses.

**Conclusions:** Our results support employing annual protective measures against SARS-CoV-2 such as administration of seasonal booster vaccines or other non-pharmaceutical interventions in a similar timeframe as those already in place for influenza prevention.

**Summary of the Main Point:** Although SARS-CoV-2 continues to cause morbidity and mortality year-round due to its high transmissibility and rapid viral evolution, our results suggest that COVID-19 activity in the United States and Europe peaks during the traditional winter viral respiratory season.

## Introduction

The Coronavirus Disease (COVID-19) pandemic has caused unprecedented worldwide morbidity, mortality, and social and economic disruption. Globally, waves of disease have primarily corresponded with the emergence of new variants of concern—which have shown increased transmissibility,^1^ improved ability to evade vaccine- or infection-induced immunity,^2^ or both. Vaccination strategies to date have struggled to keep pace, and booster doses have been deployed to bolster protection against infection and symptomatic disease and maintain peak levels of protection against severe disease throughout the pandemic.^3–7^

Nearly all respiratory viruses capable of causing human infection show distinct seasonal patterns and result in waves of illness during the winter months.^8,9^ These patterns are likely caused by a combination of host, pathogen, and environmental factors, including increased indoor activity in the winter months and seasonal temperature and humidity fluctuations known to impact viral stability outside of the host.^8,10–12^ To date, there is still speculation about whether SARS-CoV-2 currently follows—or will follow in the future—similar seasonal patterns.^13–15^ Determining this is critical for public health planning, including informing vaccine policy about the optimal timing for deploying booster doses. To help answer this urgent public health question, we evaluated whether rates of COVID-19 in the United States and Europe followed a seasonal pattern using time series models.

## Methods

### Primary analysis

Data describing daily frequencies of COVID-19 cases were obtained from the public-use Our World in Data (OWID) website. For analysis purposes, daily data from OWID were aggregated into weeks for the period between weeks ending March 3, 2020, and April 9, 2022. We calculated rates for the United States and European Union Five countries (EU5; France, Germany, Italy, Spain, and the United Kingdom) using country-specific estimated population sizes and report rates per 1,000,000 for new weekly cases during the study period.^16^ We further stratified data from the United States into the four US Census Bureau Regions (Northeast, Midwest, West, and South).^17^

For our primary analysis, we used Twitter’s time series decomposition followed by generalized extreme studentized deviate (GESD) anomaly detection to identify outlying COVID-19 rates over time.^18^ This method has been shown to reliably detect anomalies in other transmissible infectious diseases.^19–21^ This decomposition and anomaly detection method first decomposes time series data into trend, seasonal, and remainder components. The trend component is extracted using piecewise medians and median absolute deviations, which are appropriate for weekly seasonality and multimodal observed data.^18^ After decomposition, anomalies were detected using GESD, which does not require *a priori* specification of the number of outliers (as many other outlier tests do), but instead requires the specification of a range for what should be considered non-outlying data points. ^22^ We specified a 95% normal range (alpha = 0.05) with a maximum of 30% of the data to be detected as anomalous to balance the risk of false positives and negatives.

### Sensitivity analyses

To test the robustness of our findings, we performed several sensitivity analyses. First, we evaluated the impact of choosing different thresholds for the maximum amount of data to be detected as anomalous (ranging from 1–50%) compared to the base case of 30% for both the US and EU5 OWID data. To determine the impact of the data source, we repeated the primary analysis (described above) using national COVID-19 case rate data from the US Centers for Disease Control and Prevention (CDC) instead of US data derived from OWID.^23^

In addition to the methodology used in the primary analysis (i.e., Twitter’s time series decomposition with GESD anomaly detection), we also used Meta’s (previously Facebook) Prophet^24,25^ approach to decompose US time series rates and adjust for potentially confounding factors in an additive, linear time series. In the Meta Prophet model, we adjusted for the age-specific proportion of fully vaccinated individuals over time, US holidays, predominant circulating variant (omicron, delta, or alpha or wild type), and seasonality (weekly and yearly). US vaccine uptake data were obtained from US CDC.^26^ Age-group-specific vaccine uptake data were collapsed into a single value for each day by calculating a weighted average of the age-group-specific proportion of the population who was fully vaccinated (i.e., defined as two doses of mRNA vaccine or one dose of a single dose adenoviral vector vaccine) and corresponding age-group-specific population size based on 2022 United States Census Bureau estimates.^27^ Binary indicators of US holidays included were specified by the model as regressors based on calendar day(s). They included New Year’s Day, Martin Luther King Jr. Day, Washington’s Birthday, Memorial Day, Independence Day (Observed), Independence Day, Labor Day, Columbus Day, Veterans Day, Thanksgiving, Christmas Day, Christmas Day (Observed), and New Year’s Day (Observed). We also added holiday effects for two weeks of an annual spring break (Mar 16, 2020 – Mar 27, 2020; Mar 15, 2021 – Mar 26, 2021; and Mar 14, 2022 – Mar 25, 2022) and extra days before Christmas Day through New Year’s Day to account for potential significant holiday travel effects (Dec 21, 2020 – Jan 3, 2021; and Dec 20, 2021 – Jan 2, 2022). SARS-CoV-2 variant frequencies by date for the United States were obtained from covariants.org.^28^ Variant frequencies were summed by week to match COVID-19 case data. Primary circulating variant was defined as the most frequent variant identified each week and was categorized into three groups: wild type/Wuhan or alpha (Mar 9, 2020 – Apr 4, 2021), delta (Apr 5, 2021 – Dec 13, 2021), or omicron (Dec 14, 2021 – Apr 9, 2022). In the Prophet model, weekly and yearly seasonality were set to automatic detection, while daily seasonality was turned off as we only had daily reports that were not time-stamped. Uncertainty interval widths were set to 95%, with 2,000 uncertainty samples used to compute the intervals. Markov chain Monte Carlo simulations (n=1,000) were used to calculate uncertainty intervals for the Prophet time series decomposed components.

Finally, as a confirmatory sensitivity analysis, we applied both the anomaly detection and Prophet modeling approaches to US influenza data (rather than COVID-19 data) to assess the model would predict well-known seasonal patterns for influenza. Influenza case counts for the United States were obtained from US CDC FluView and included weekly case counts of influenza viruses reported to CDC through the National Respiratory and Enteric Virus Surveillance System at the national level.^29^ Total influenza cases was defined as the sum of all subtypes reported to clinical and public health laboratories from Mar 2, 2014, through Apr 5, 2020. We excluded 2020/2021 and 2021/2022 influenza seasons due to the confounding effects of COVID-19 precautions on reducing the influenza burden. ^30^

For anomaly detection, a base case of 30% maximum anomalies was used, and for Prophet modeling, US holidays, the primary circulating variant of the week, and the weekly population weighted mean of percent vaccine uptake were used for adjustment as indicated above for COVID-19 data. R version 4.1.2 (R Foundation for Statistical Computing, Vienna, Austria) was used for all analyses.

## Results

Anomaly plots detected COVID-19 rates that were higher than expected between November and March each year in the United States (**Figure 1A**) and EU5 (**Figure 1B**). In the United States, trends were similar for the North, Midwest, and West US Census Bureau Regions, with seasonal spikes in rates observed between November and February (**Figures 2A-2C**). In the Southern Region, in addition to seasonal peaks in the fall/winter, a second peak in the late summer of 2021 was identified as anomalous in Aug/Sep 2021, which returned to normal by Oct 2021 (**Figure 2D**). This peak was not identified in the US Southern Region in 2020.

**Figure 1:**
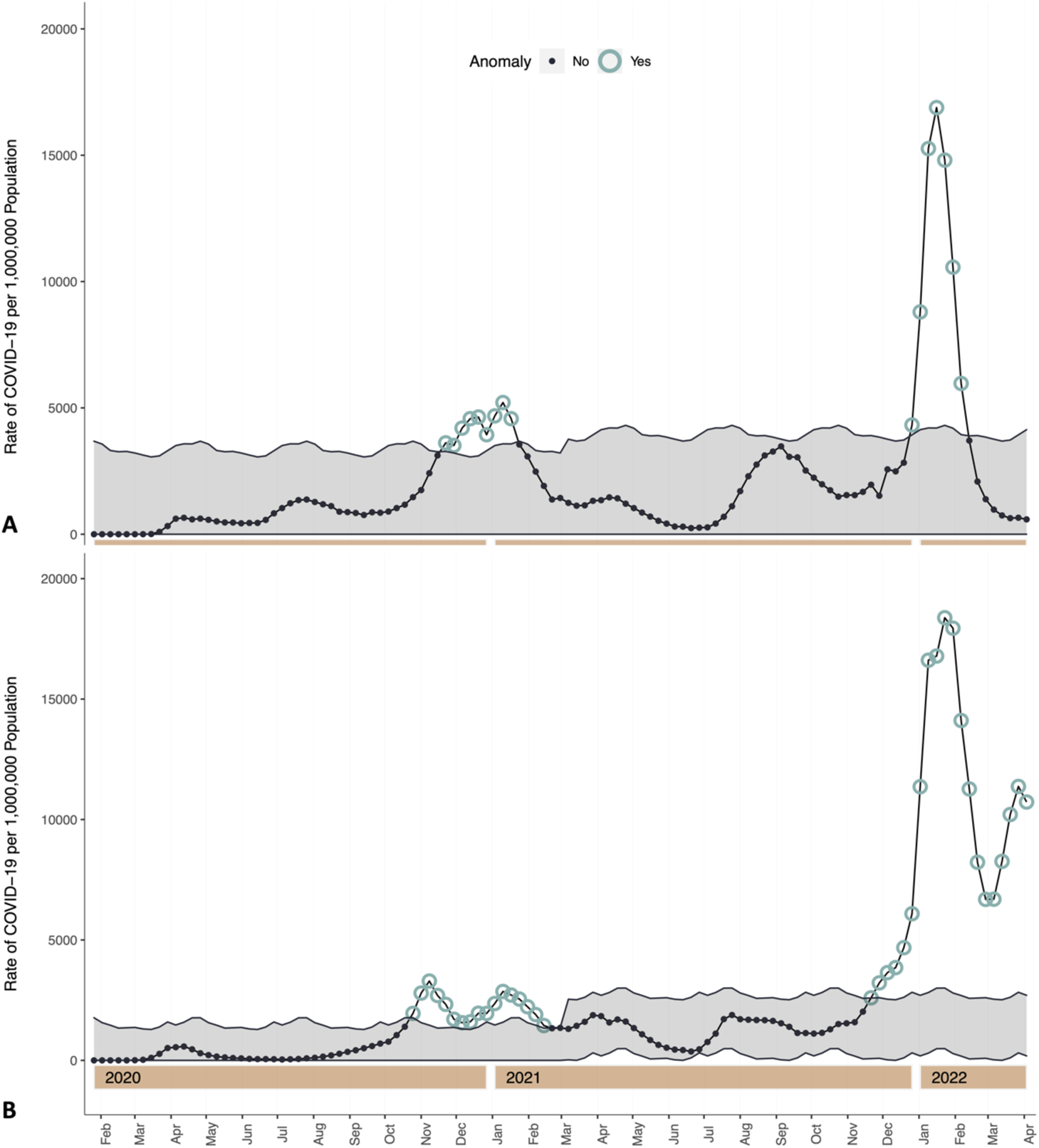
Twitter time series decomposition with generalized extreme studentized deviate (GESD) anomaly detection of COVID-19 rates, Mar 7, 2020 – Apr 9, 2022. **Panel A**: United States Our World in Data; **Panel B**: EU5: Italy, Germany, France, Spain, and the United Kingdom, Our World in Data. Shaded areas represent the normal range of data points.

**Figure 2:**
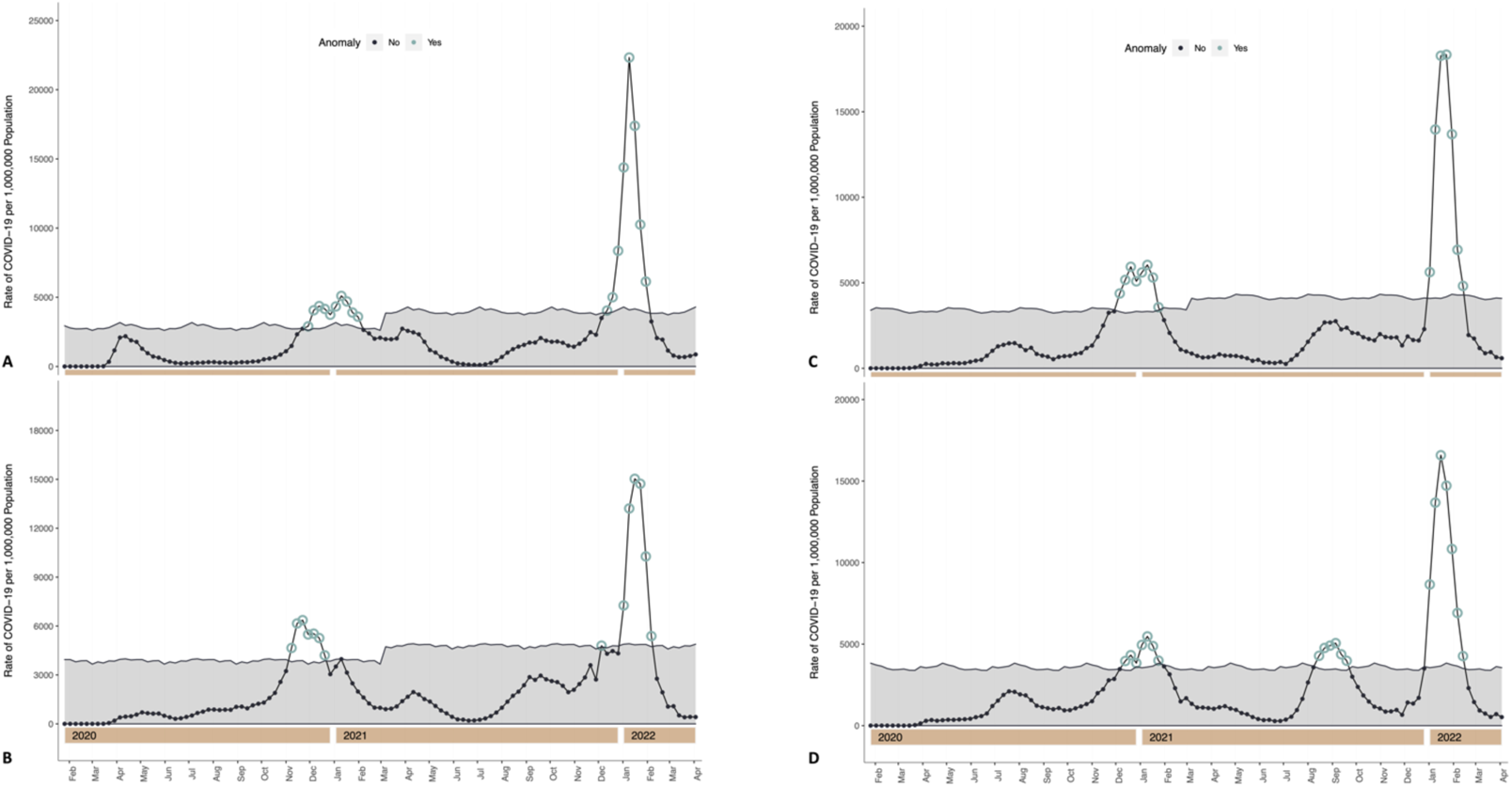
Twitter time series decomposition with generalized extreme studentized deviate (GESD) anomaly detection of COVID-19 rates, Mar 7, 2020 – Apr 9, 2022, United States, by Census Region. **Panel A,** Northeast Region; **Panel B**, Midwest Region; **Panel C**, West Region; **Panel D**, South Region. Data Source: Our World in Data

Sensitivity analyses confirmed that the threshold for percent of observations detected as anomalous (supplementary appendix, **Figures s1 – s12**) had little impact on observed seasonal trends. In addition, seasonal US COVID-19 trends were similar if US CDC data were used instead of data from OWID (**Figure s13**). Sensitivity analyses using Meta’s Prophet model also showed a strong annual seasonal component for COVID-19 case rates from approximately December through February in the United States (**Figure 3A**), similar to the primary analysis. During the study period, the general trend of COVID-19 was increasing (**Figure 3B**), although with wide uncertainty intervals. Several strong holiday effects were identified near Christmas and New Year’s, spring break, and Independence Day (**Figure 3C**).

**Figure 3:**
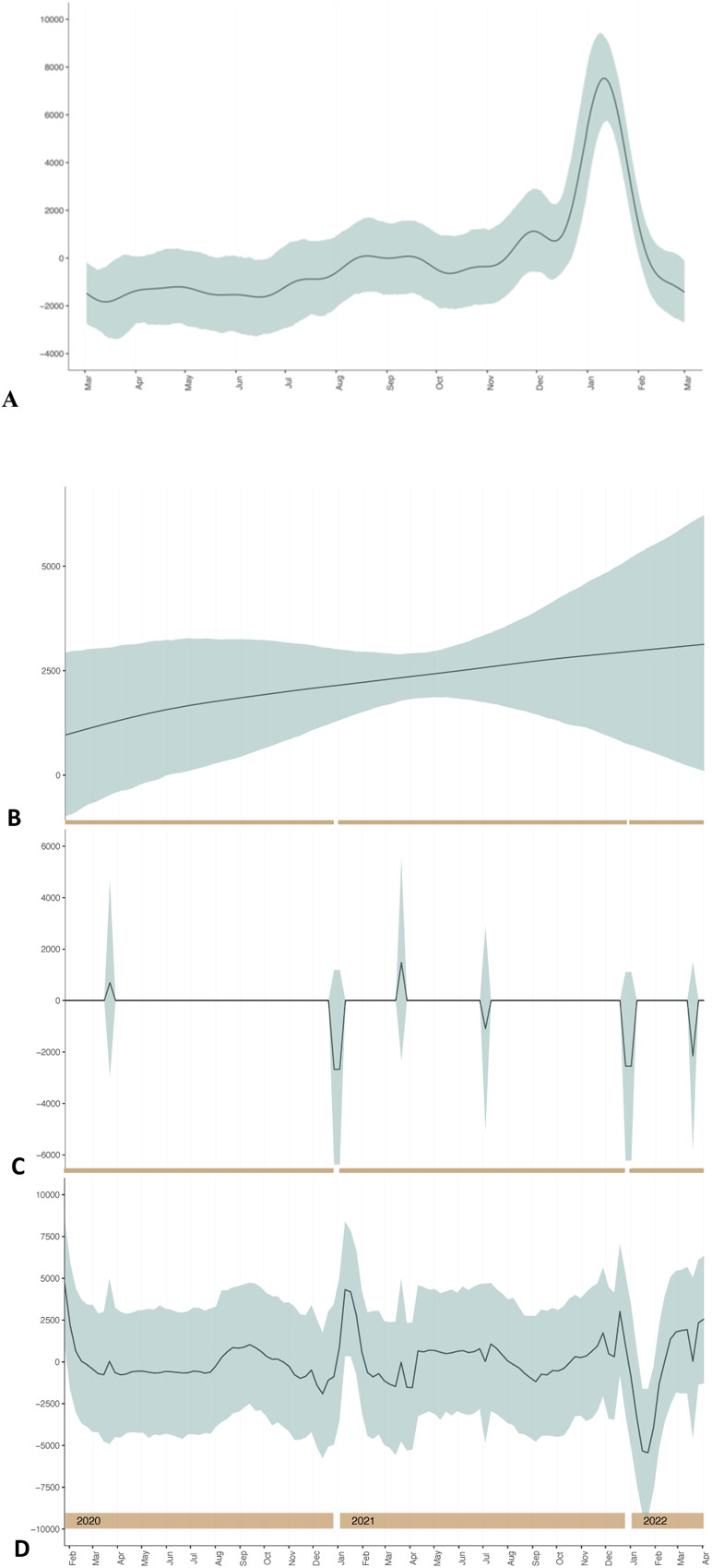
COVID-19 rates from the Prophet decomposed model with Markov chain Monte Carlo Simulated 95% uncertainty intervals, Mar 7, 2020 – Apr 9, 2022, adjusted for US holidays, dominant variant, and age-weighted vaccine uptake, United States. **Panel A**: Annual seasonal component aggregated over study period; **Panel B**, Trend Component; **Panel C**, Holiday Component; **Panel D**, Residual Component. Data Source: Our World in Data

Anomaly detection analyses using the same model specifications described for rates of COVID-19 also accurately predicted seasonal spikes of influenza between November and April over six US influenza seasons (**Figure 4**), consistent with current knowledge about annual influenza patterns in the United States.^31^ Further, Prophet modeling of influenza data showed remarkably similar trends as for COVID-19 (**Figures s14 – s17**), with slightly more residual variation, likely due to fewer regressors added to the model compared to COVID-19 data.

**Figure 4:**
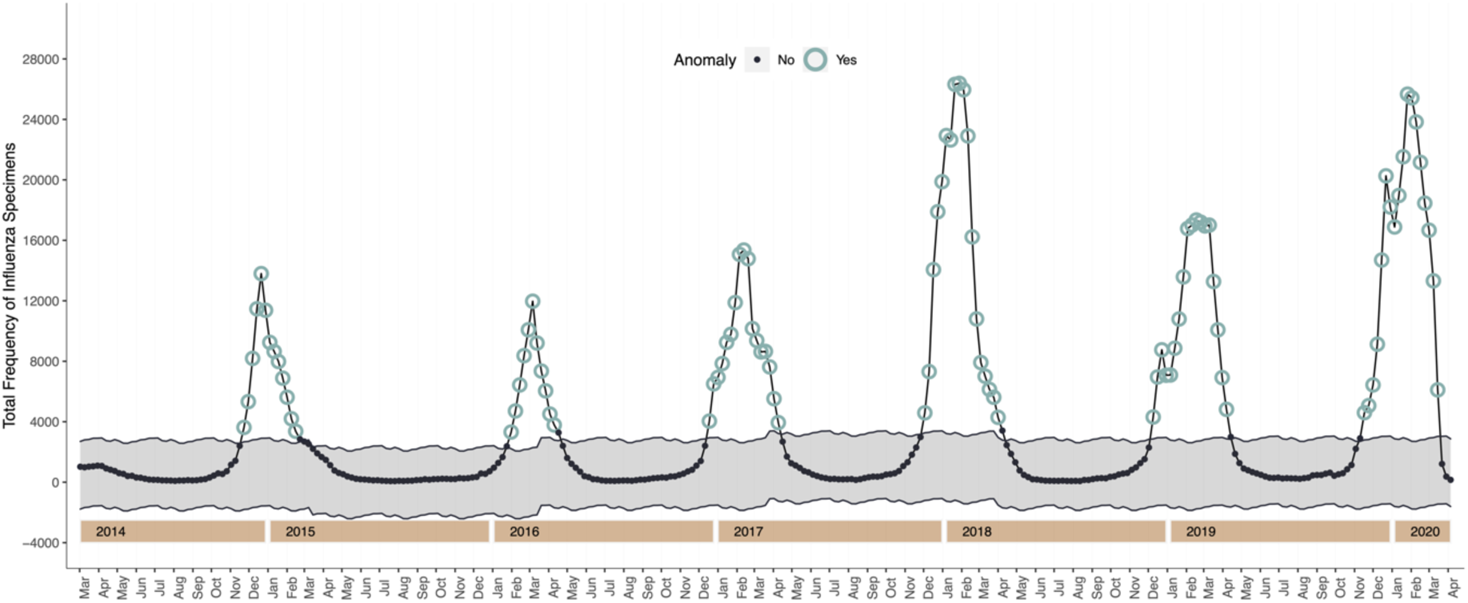
Twitter time series decomposition with generalized extreme studentized deviate (GESD) anomaly detection of Influenza cases, Mar 1, 2014 – Apr 10, 2020. Shaded areas represent the normal range of data points. Data Source: United States Centers for Disease Control and Prevention FluView

## Discussion

Although SARS-CoV-2 continues to circulate and cause disease year-round, we identified clear seasonal trends in rates of COVID-19 in the United States and Europe. Since the beginning of the pandemic, COVID-19 rates in the United States and EU5 have spiked in the months of December through March—the same months typical of seasonal respiratory virus epidemics in the northern hemisphere.^31^ These results are consistent with other coronaviruses that exhibit largely seasonal patterns^32^ and mathematical simulations of COVID-19 disease activity.^13,14^ There are many possible reasons for seasonality of respiratory viruses, including climate-related changes in viral transmissibility, modified host factors (e.g., waning of infection- or vaccine-induced immunity), and changes in human behavior during the winter months.^8,15^ Regardless of the mechanisms, knowledge of pathogen seasonality is imperative for instituting targeted interventions to decrease disease burden.

Accordingly, our findings have important vaccine policy implications. Booster doses of COVID-19 vaccines administered before the winter months will likely have the most significant public health impact on COVID-19 disease burden. This is analogous to providing influenza vaccine before peak flu activity each year. Because SARS-CoV-2 is considerably more transmissible than influenza and other seasonal respiratory viruses, it remains possible that year-round SARS-CoV-2 activity will remain elevated compared to other pathogens. Providing more than one booster dose of COVID-19 vaccines each year, however, has proven programmatically challenging, and concerns regarding “booster fatigue” are increasing.^33,34^ Thus, timing the administration of an annual COVID-19 vaccine, and thus peak vaccine protection, with the likely timing of peak COVID-19 disease activity (i.e., the winter viral respiratory season based on our results) may be the most prudent approach in the near term. Despite evidence that protection provided by current mRNA COVID-19 vaccines wanes significantly against omicron infection and symptomatic disease after only 3 to 4 months—even after a booster^4,7,35^—this short-term protection could still provide meaningful defense against SARS-CoV-2 infection if deployed just before seasonal waves that last 3 to 4 months on average. Moreover, it remains unknown whether variant-adapted vaccines may improve durability of protection against infection and symptomatic disease for even longer than current wild-type formulations. Whether additional boosters at a frequency greater than once annually are needed for some high-risk groups will likely be a careful balance between epidemiological, benefit-risk, and programmatic considerations moving forward.

There has been much debate about whether the goal of vaccination programs should be only to prevent severe disease or if it should include preventing infection and reducing transmission.^36^ We now know that vaccination alone is unlikely to lead to the eradication or elimination of SARS-CoV-2. However, deploying vaccines on schedule that times peak protection to correspond with peak disease activity can still have a meaningful impact on flattening future waves of infection and disease in addition to ensuring protection against severe illness is maintained year-over-year. Lessening the burden of SARS-CoV-2 infection remains an important goal and corresponds with fewer long-term consequences of infection such as post-acute sequelae^37–39^ and other disruptive societal and economic consequences.^40^

A more nuanced finding from our study was that, in the United States, southern States might be experiencing second summer waves, as has been previously suggested.^41^ There could be many reasons behind this additional summer wave. Some possible explanations include lower vaccination rates or use of nonpharmaceutical interventions in southern US States compared to other regions,^42^ changes in behavioral patterns including end-of-summer vacations to southern States, or increased indoor gathering due to excessive heat during this time in the US south. Visually, there were slight increases in rates within other US Census regions during this time, however, they were within the expected variability of case rates. It is reasonable to assume that there would have been higher rates during these months in all regions had adequate protections (e.g., vaccination, nonpharmaceutical interventions) been less prevalent.

In sensitivity analyses, our methodology accurately detected anomalous rates of influenza virus infection in the United States, underscoring the utility of anomaly detection for detecting seasonal patterns in common respiratory viruses. Although the ‘influenza season’ appeared to be slightly longer than the ‘COVID-19 season,’ this could be due to the additional prevention measures taken for COVID-19, including masking and social distancing. These interventions are highly effective for influenza, as evidenced by the near disappearance of influenza infection during the height of the COVID-19 pandemic in the United States.^30^

Our results have limitations. First, we could not account for potential underreporting of cases, which may have a large effect more recently with increases in at-home SARS-CoV-2 testing that may not be reported.^43^ Further, statistical modeling may not fully reflect the intricacies of preventing transmissible infectious diseases, such as the impact of waning immunity or changes in testing, nonpharmaceutical interventions, or healthcare-seeking behavior over time. Despite this, our sensitivity analysis using Meta’s Prophet model, which controlled for vaccination coverage over time, holidays, and the dominant SARS-CoV-2 variant each week, also showed a strong annual seasonal component for rates of COVID-19. Moreover, we found seasonal patterns over the duration of the entire pandemic despite changes in testing and mitigation behaviors over time. Although the pandemic is in its third year, the amount of data available for forecasting was limited compared to other common seasonal viruses. Because of this, forecasts may change rapidly in the months or years ahead. Another limitation is that our findings are not generalizable beyond the United States and Europe, and more research is needed to understand if seasonal patterns in SARS-CoV-2 activity are seen in the Southern Hemisphere or Asia-Pacific regions. Finally, with SARS-CoV-2, there is always the potential for new variants to emerge that could meaningfully escape prior vaccine- or infection-induced immunity and cause epidemics outside of regular seasonal patterns. Thus, the public health community should continue to plan and maintain capability for this possibility.

Although SARS-CoV-2 continues to cause morbidity and mortality year-round due to its high transmissibility and continued rapid viral evolution, our results suggest that COVID-19 activity in the United States and Europe peaks during the traditional winter viral respiratory season. Thus, employing annual protective measures against SARS-CoV-2 such as administering seasonal booster vaccines or other non-pharmaceutical interventions in a similar timeframe as those already in place for influenza prevention (i.e., beginning in early autumn) is a prudent strategy to stay ahead of likely forthcoming seasonal waves of COVID-19. Additional confirmatory studies including those conducted in the Southern Hemisphere and other regions outside of the United States and Europe, however, are needed.

## Data Availability

All data produced are available online from Our World In Data and the US Centers for Disease Control and Prevention COVID-19 Data Tracker and FluView websites.

## Author Contributions

Dr. Wiemken had full access to all the data in the study and took responsibility for the integrity and data analysis accuracy. All authors had full access to all data and accept responsibility for submitting for publication.

*Concept and design:* All Authors.

*Accessed and verified data and analysis: TLW, FK Interpretation of data:* All authors.

*Drafting of the manuscript:* TLW, JMM

*Critical revision of the manuscript for important intellectual content:* All Authors

*Obtained funding:* N/A.

*Administrative, technical, or material support:* All Authors

*Supervision:* JMM

## Data Sharing

All data utilized in this study are open and available to the public free of charge, as referenced.

## Supplementary Appendix

**Figure s1.**
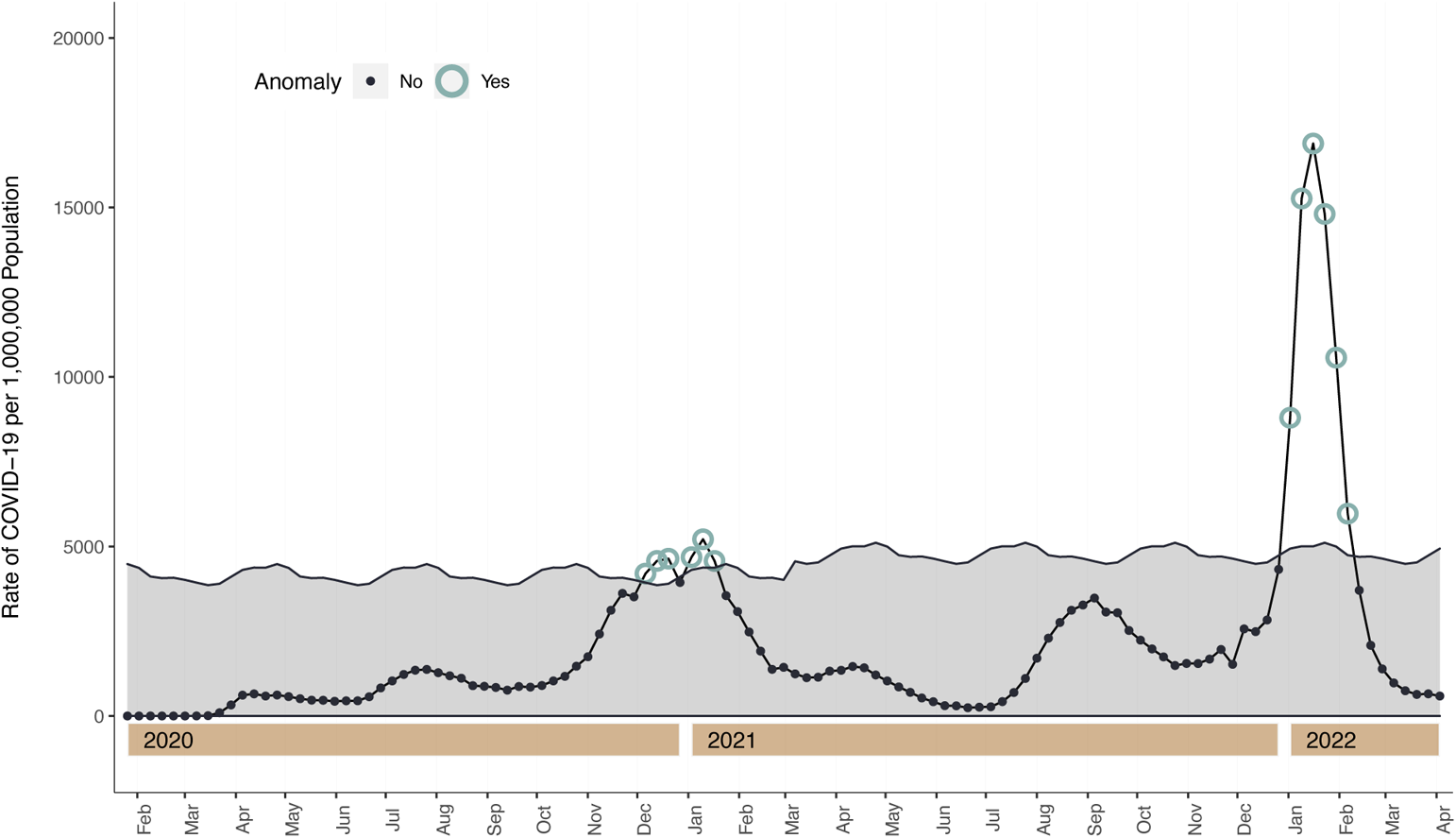
Twitter time series decomposition with generalized extreme studentized deviate (GESD) anomaly detection of US COVID-19 case rates, Mar 7, 2020 – Apr 9, 2022, with 1% maximum anomalies, Data Source: OWID

**Figure s2.**
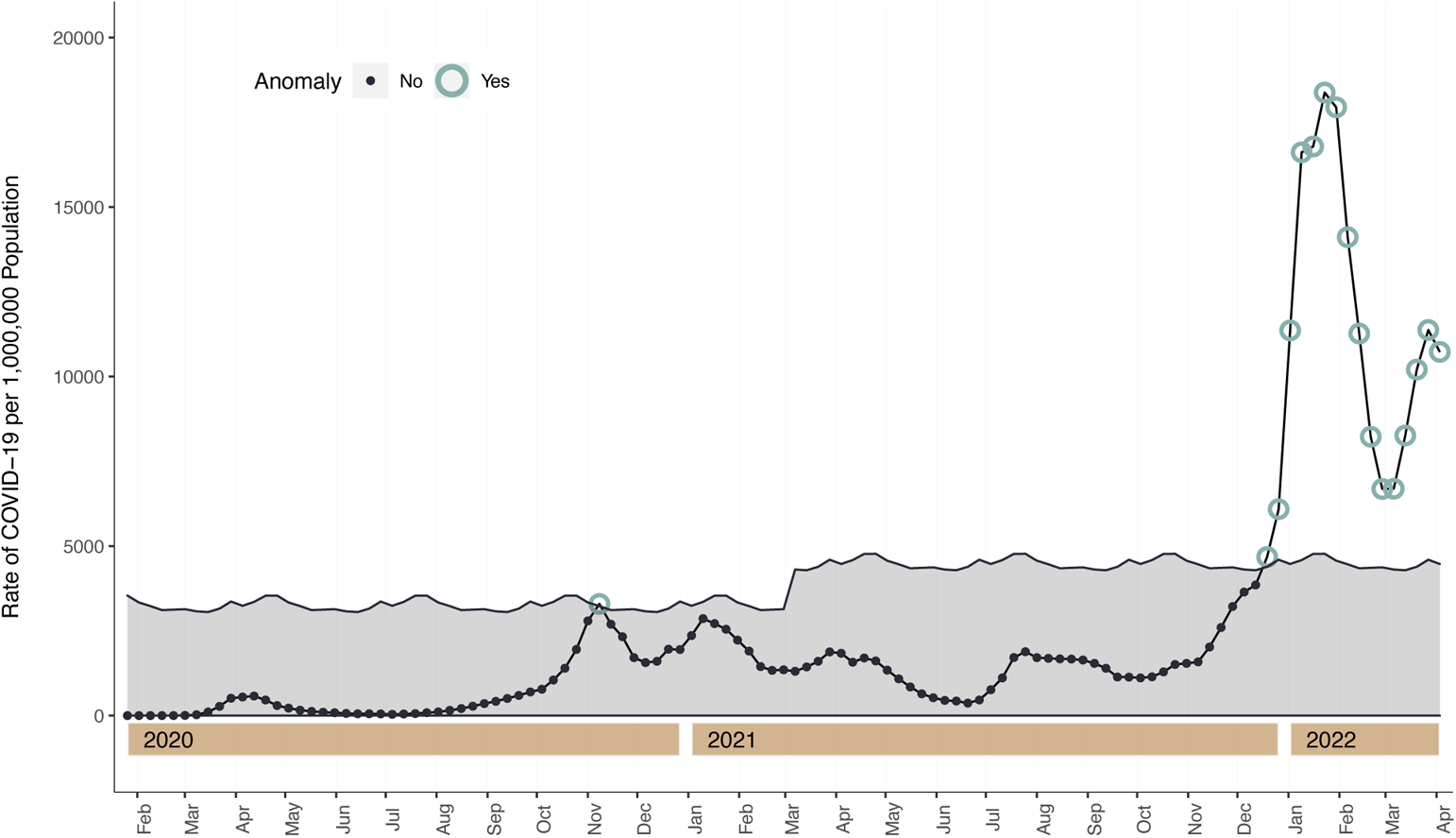
Twitter time series decomposition with generalized extreme studentized deviate (GESD) anomaly detection of EU-5 Country COVID-19 case rates, Mar 7, 2020 – Apr 9, 2022, with 1% maximum anomalies, Data Source: OWID

**Figure s3.**
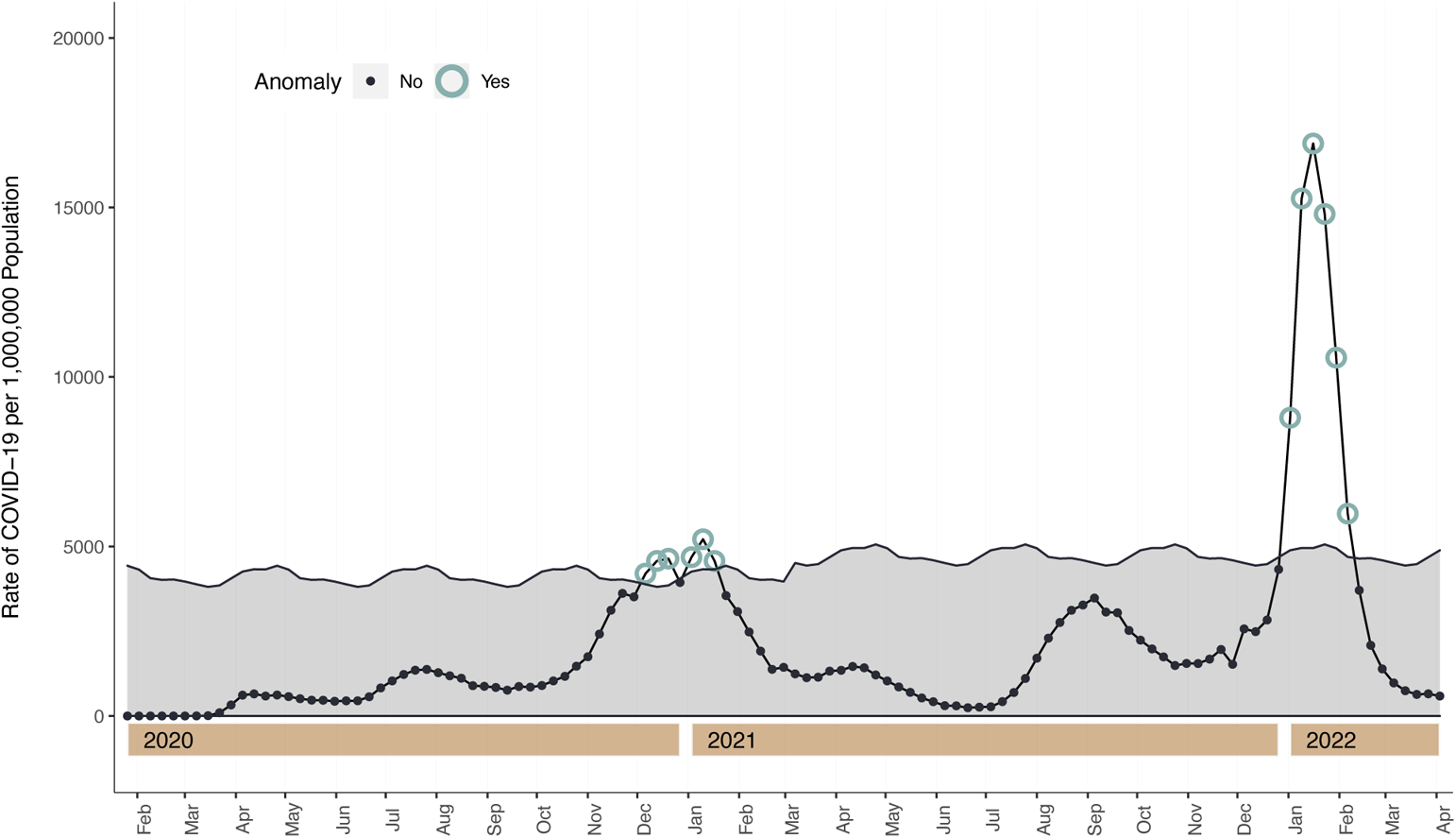
Twitter time series decomposition with generalized extreme studentized deviate (GESD) anomaly detection of US COVID-19 case rates, Mar 7, 2020 – Apr 9, 2022, with 5% maximum anomalies, Data Source: OWID

**Figure s4.**
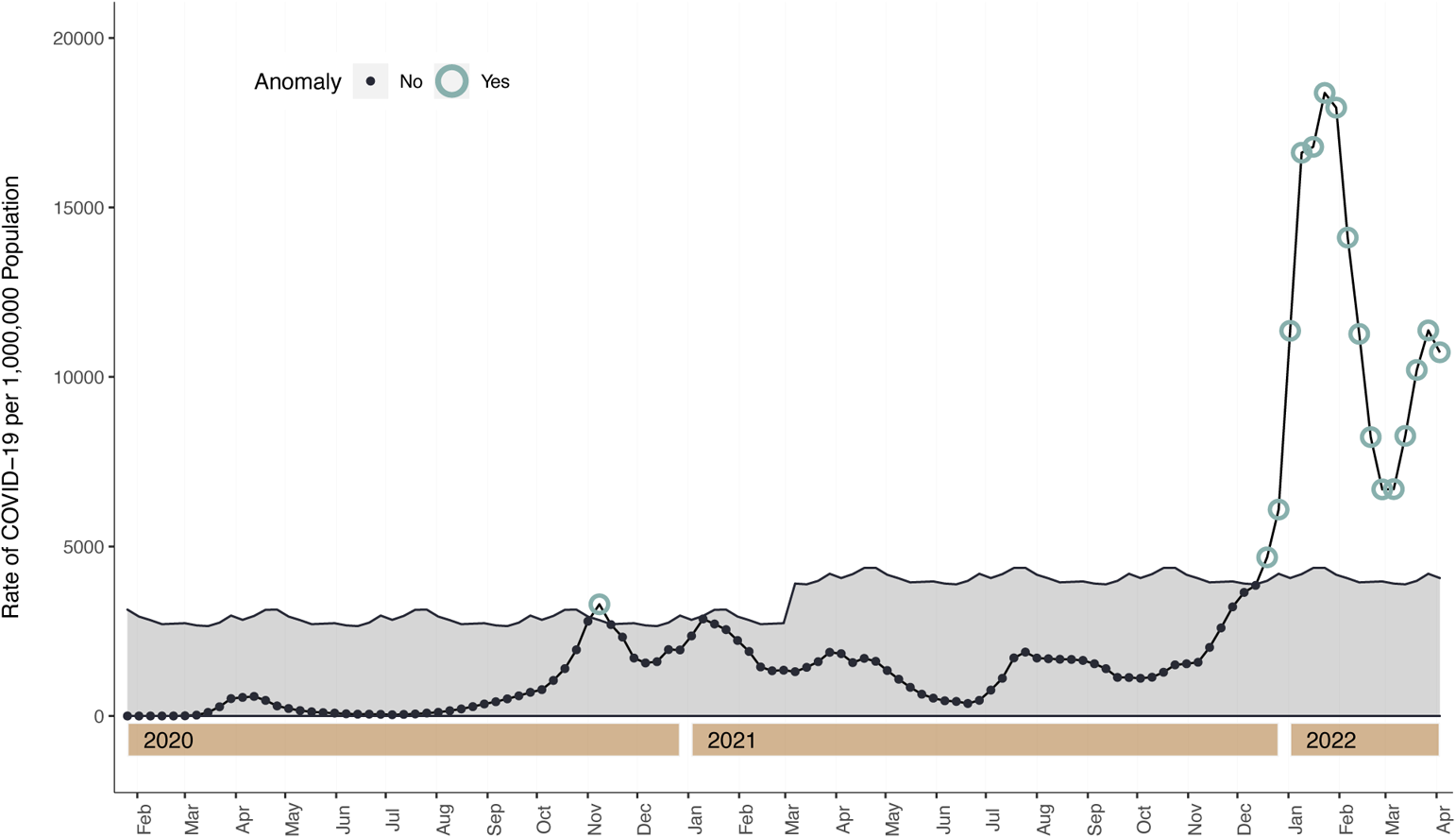
Twitter time series decomposition with generalized extreme studentized deviate (GESD) anomaly detection of EU-5 Country COVID-19 case rates, Mar 7, 2020 – Apr 9, 2022, with 5% maximum anomalies, Data Source: OWID

**Figure s5.**
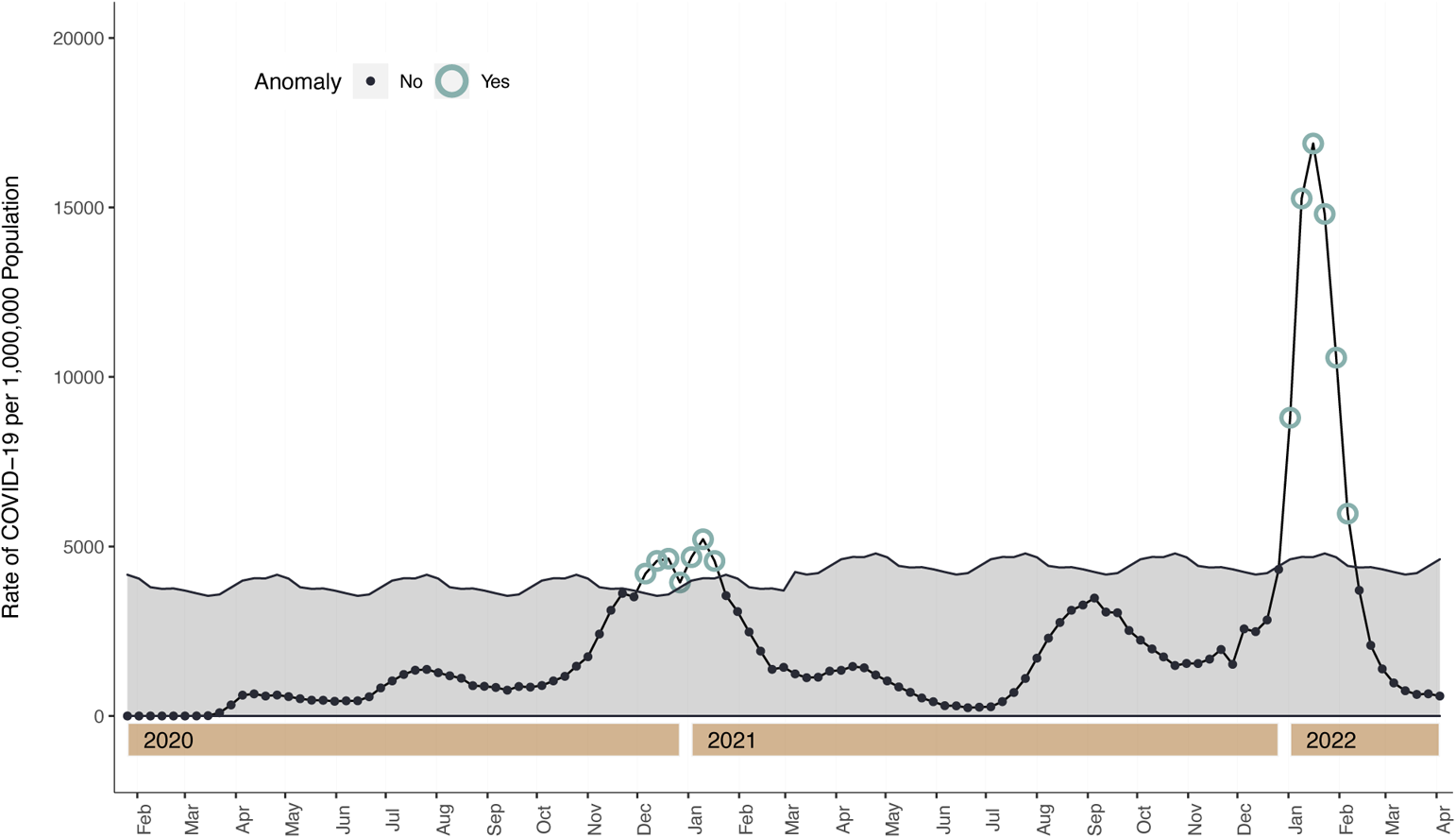
Twitter time series decomposition with generalized extreme studentized deviate (GESD) anomaly detection of US COVID-19 case rates, Mar 7, 2020 – Apr 9, 2022, with 10% maximum anomalies, Data Source: OWID

**Figure s6.**
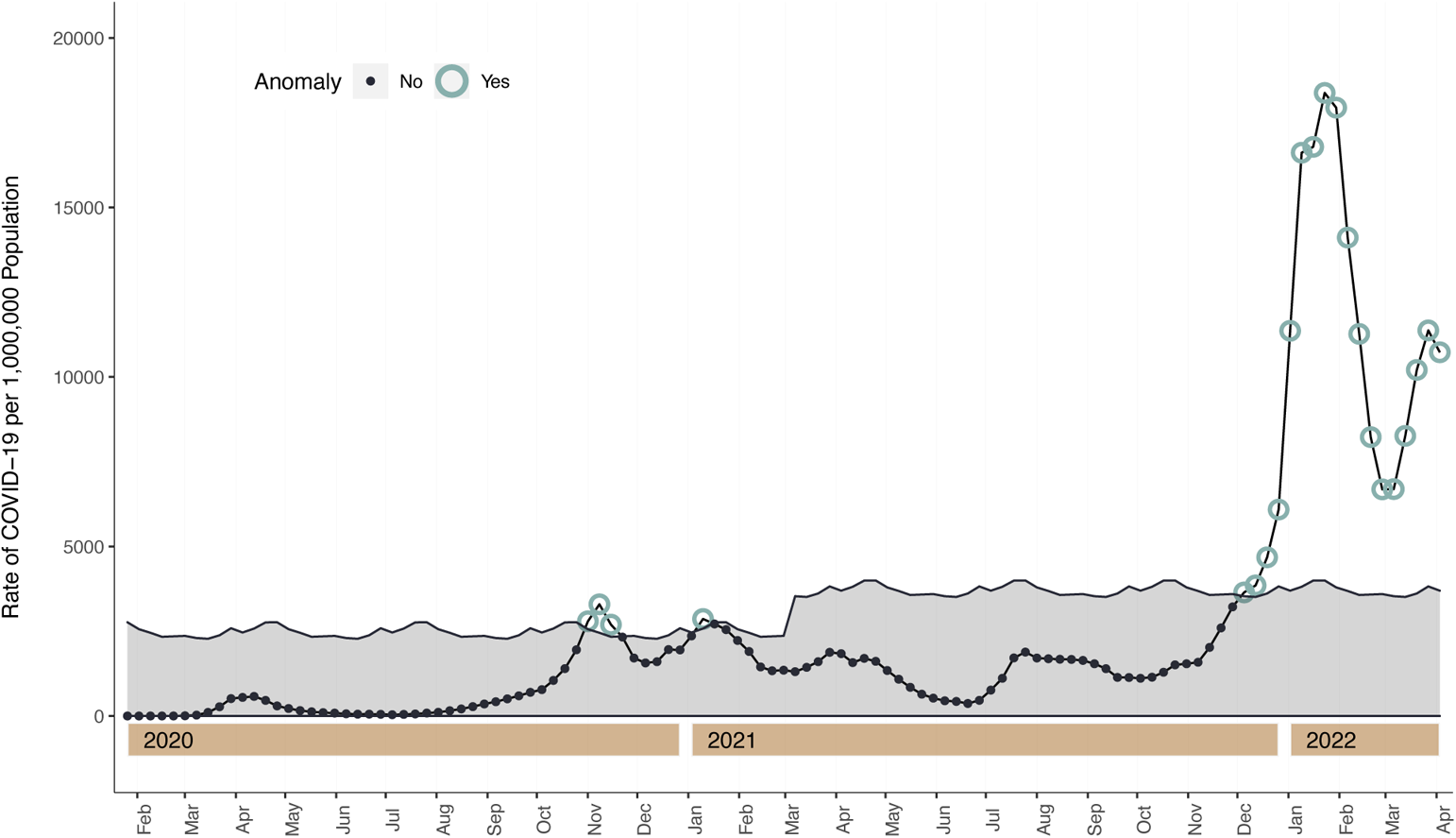
Twitter time series decomposition with generalized extreme studentized deviate (GESD) anomaly detection of EU-5 Country COVID-19 case rates, Mar 7, 2020 – Apr 9, 2022, with 10% maximum anomalies, Data Source: OWID

**Figure s7.**
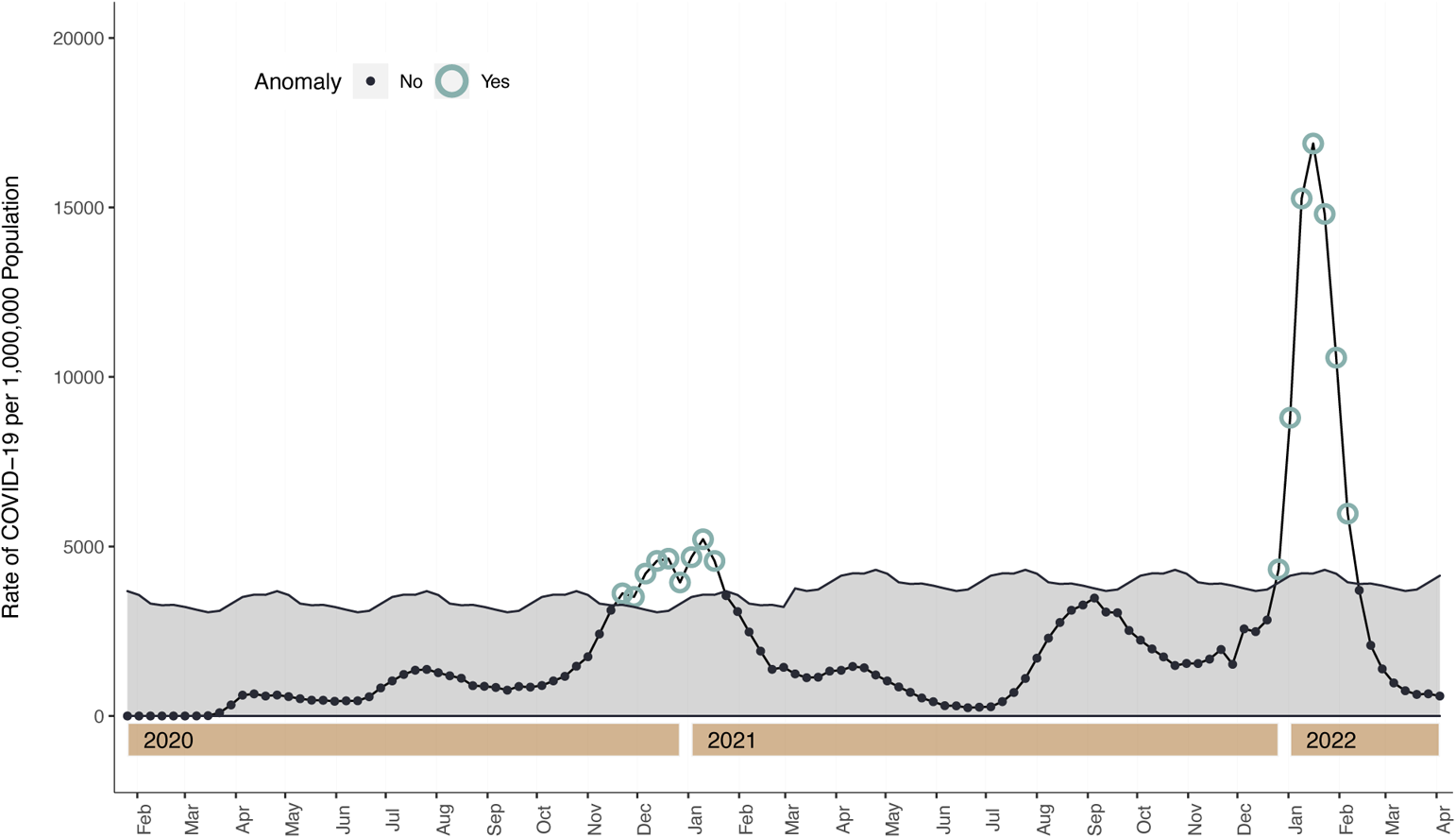
Twitter time series decomposition with generalized extreme studentized deviate (GESD) anomaly detection of US COVID-19 case rates, Mar 7, 2020 – Apr 9, 2022, with 20% maximum anomalies, Data Source: OWID

**Figure s8.**
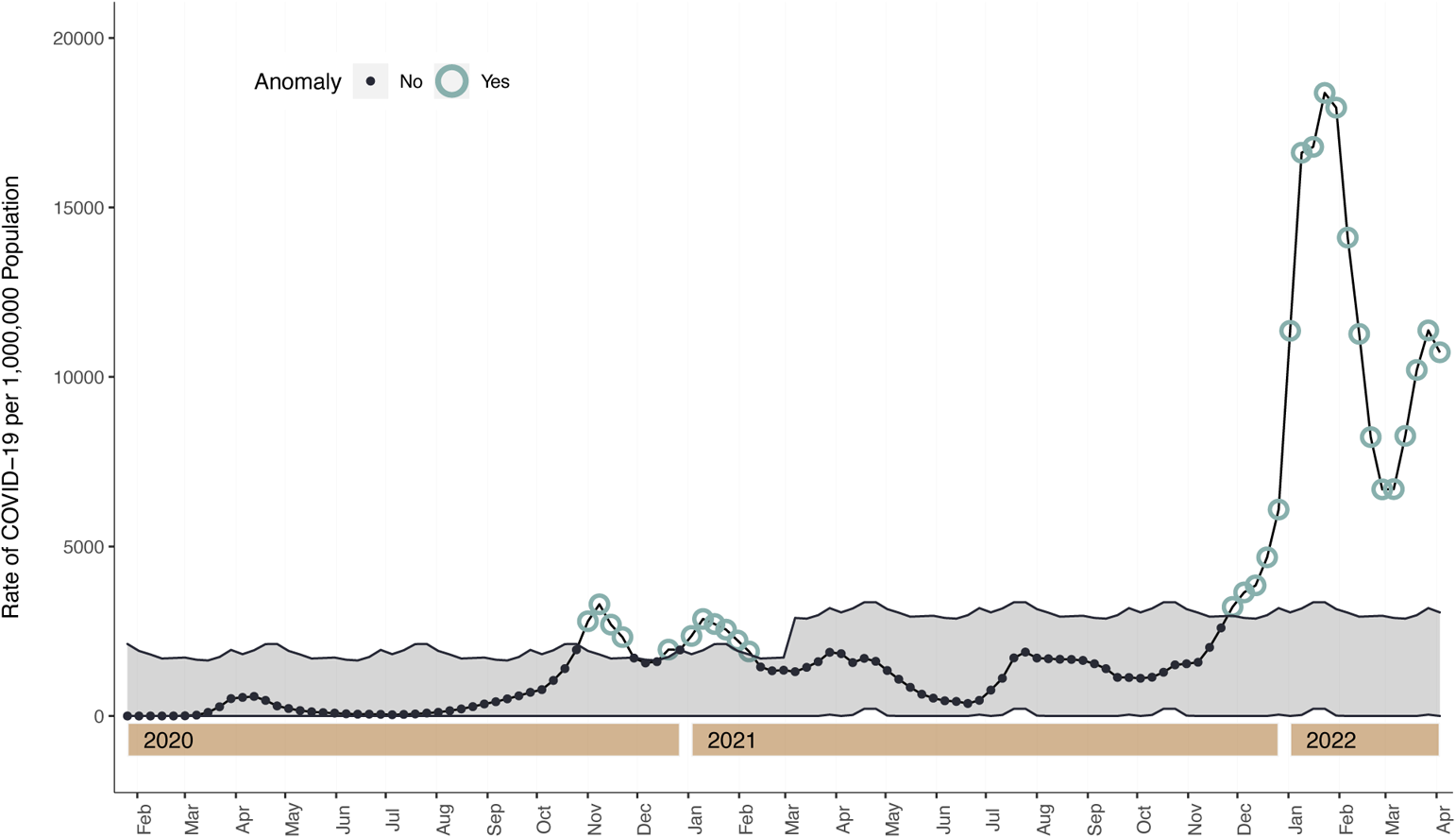
Twitter time series decomposition with generalized extreme studentized deviate (GESD) anomaly detection of EU-5 Country COVID-19 case rates, Mar 7, 2020 – Apr 9, 2022, with 20% maximum anomalies, Data Source: OWID

**Figure s9.**
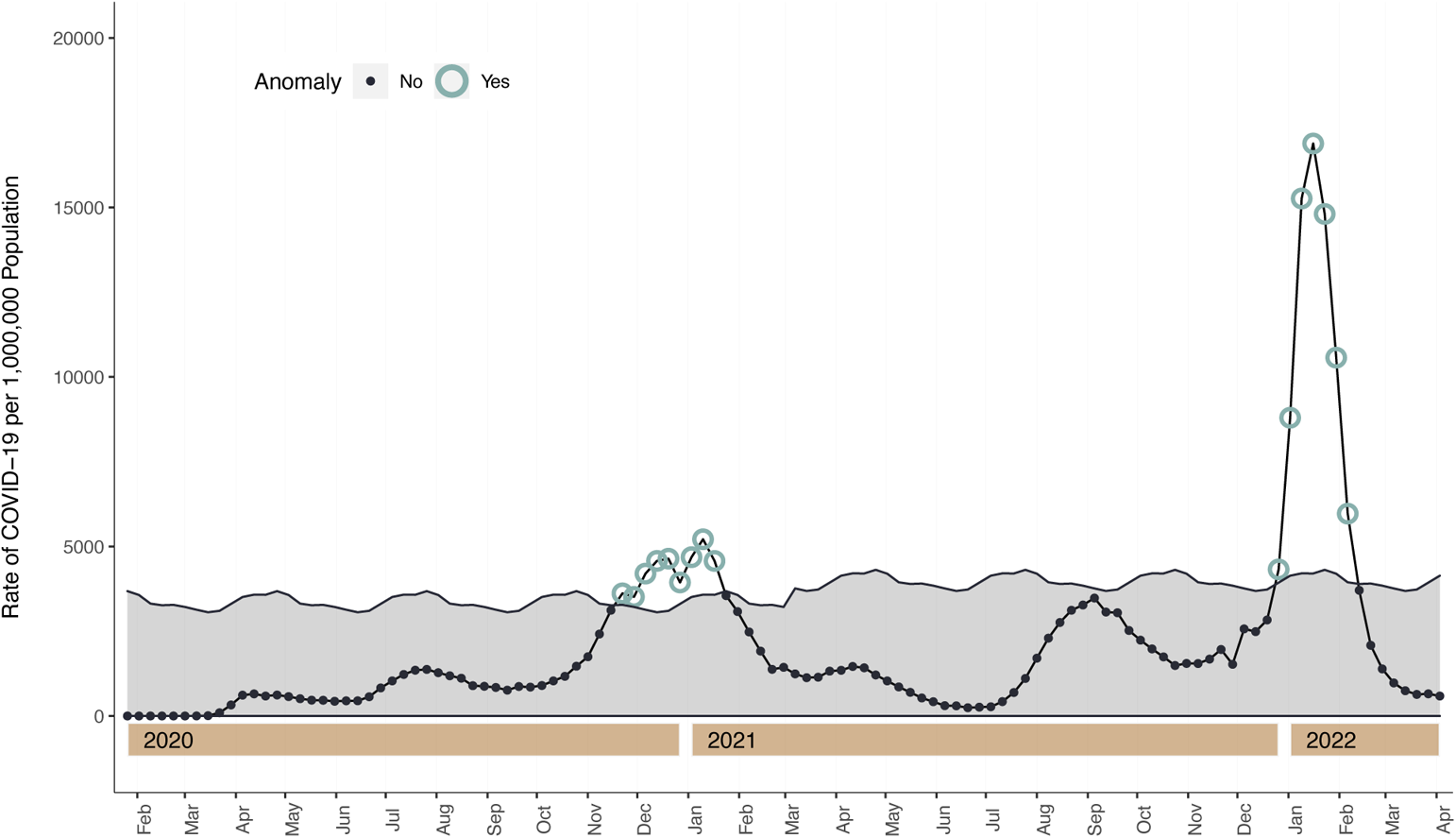
Twitter time series decomposition with generalized extreme studentized deviate (GESD) anomaly detection of US COVID-19 case rates, Mar 7, 2020 – Apr 9, 2022, with 40% maximum anomalies, Data Source: OWID

**Figure s10.**
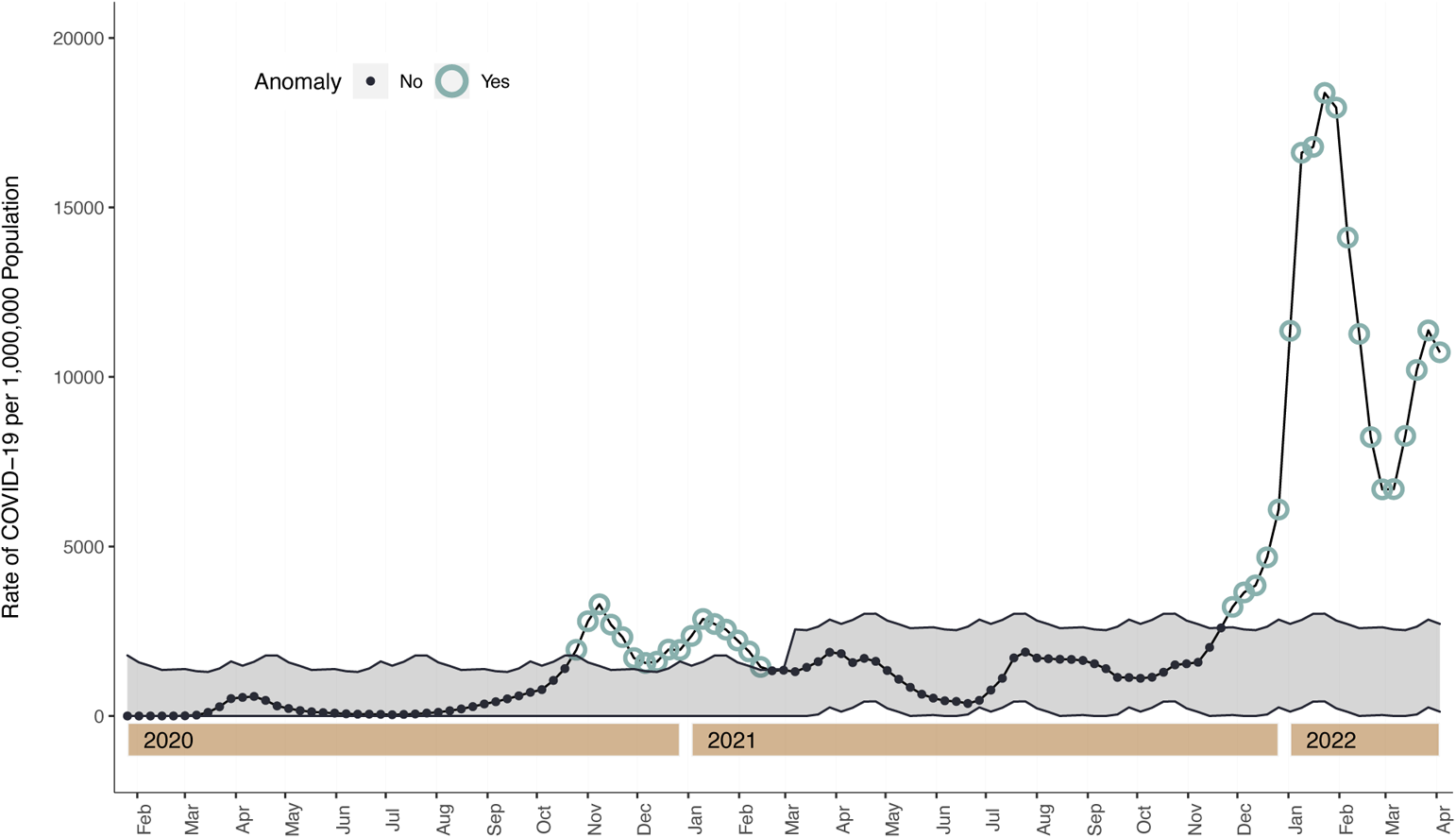
Twitter time series decomposition with generalized extreme studentized deviate (GESD) anomaly detection of EU-5 Country COVID-19 case rates, Mar 7, 2020 – Apr 9, 2022, with 40% maximum anomalies, Data Source: OWID

**Figure s11.**
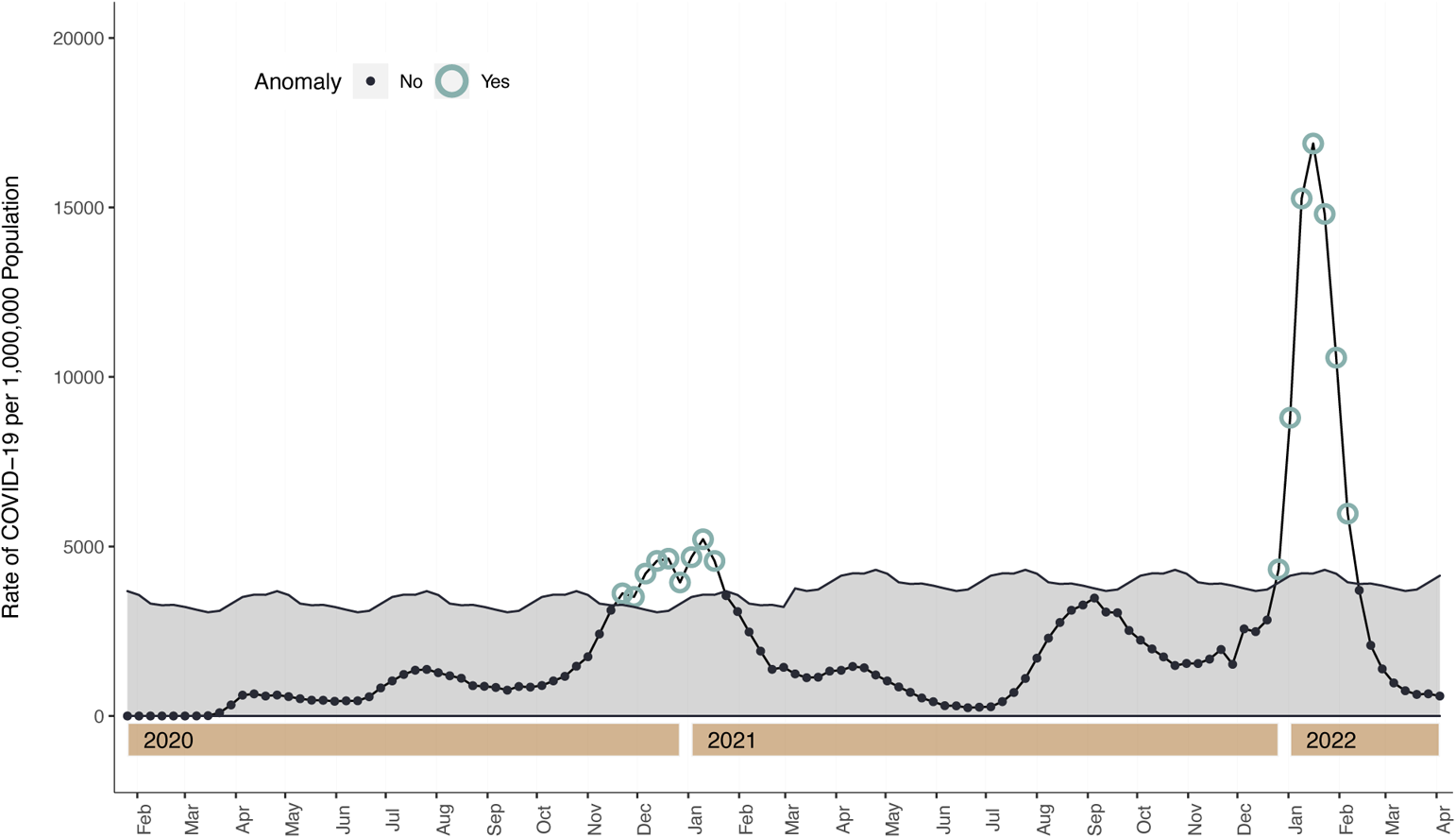
Twitter time series decomposition with generalized extreme studentized deviate (GESD) anomaly detection of US COVID-19 case rates, Mar 7, 2020 – Apr 9, 2022, with 50% maximum anomalies, Data Source: OWID

**Figure s12.**
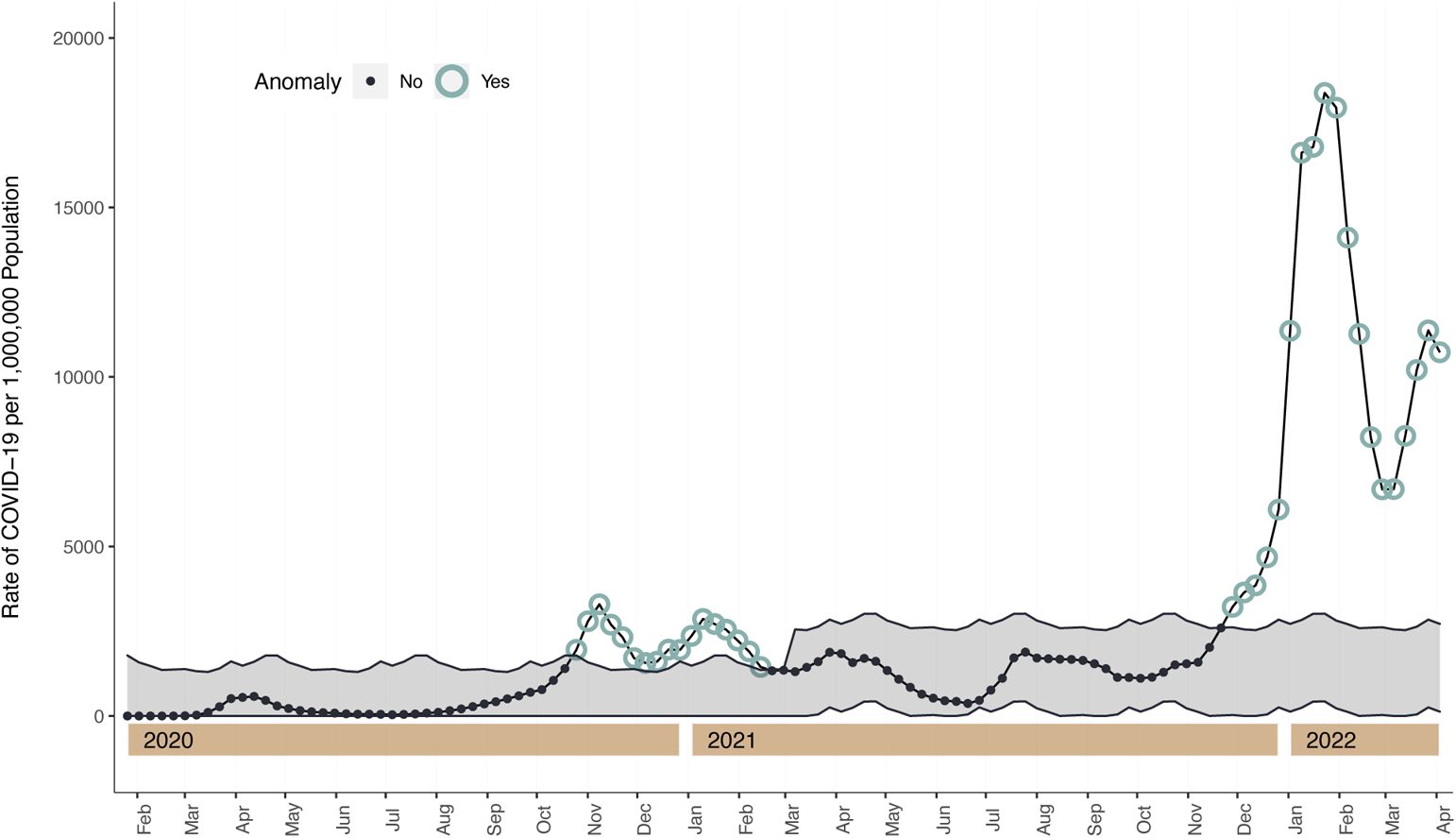
Twitter time series decomposition with generalized extreme studentized deviate (GESD) anomaly detection of EU-5 Country COVID-19 case rates, Mar 7, 2020 – Apr 9, 2022, with 50% maximum anomalies, Data Source: OWID

**Figure s13.**
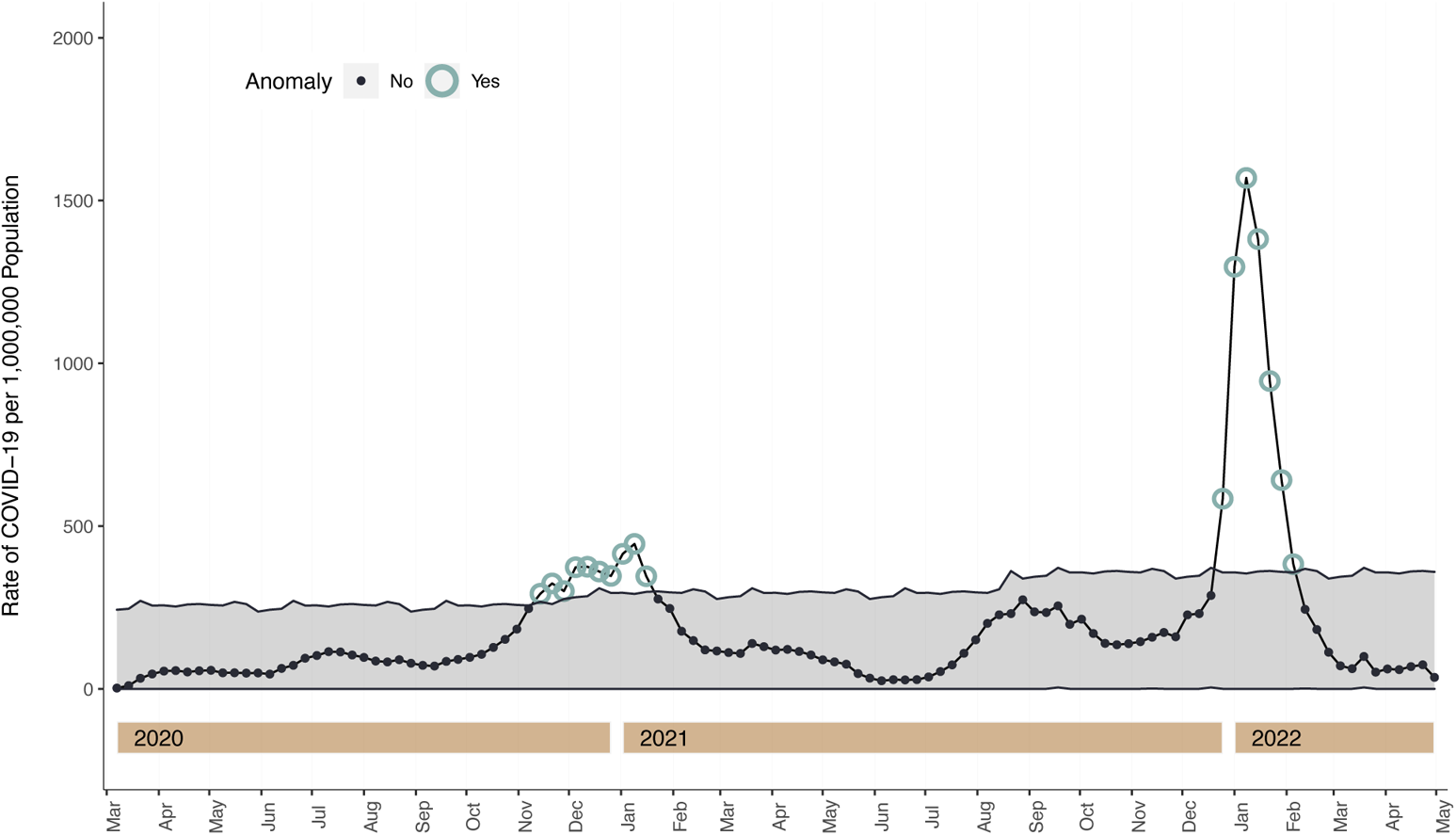
Twitter time series decomposition with generalized extreme studentized deviate (GESD) anomaly detection of US COVID-19 case rates, Mar 7, 2020 – Apr 9, 2022, with 30% maximum anomalies, Data Source: US CDC

**Figure s14.**
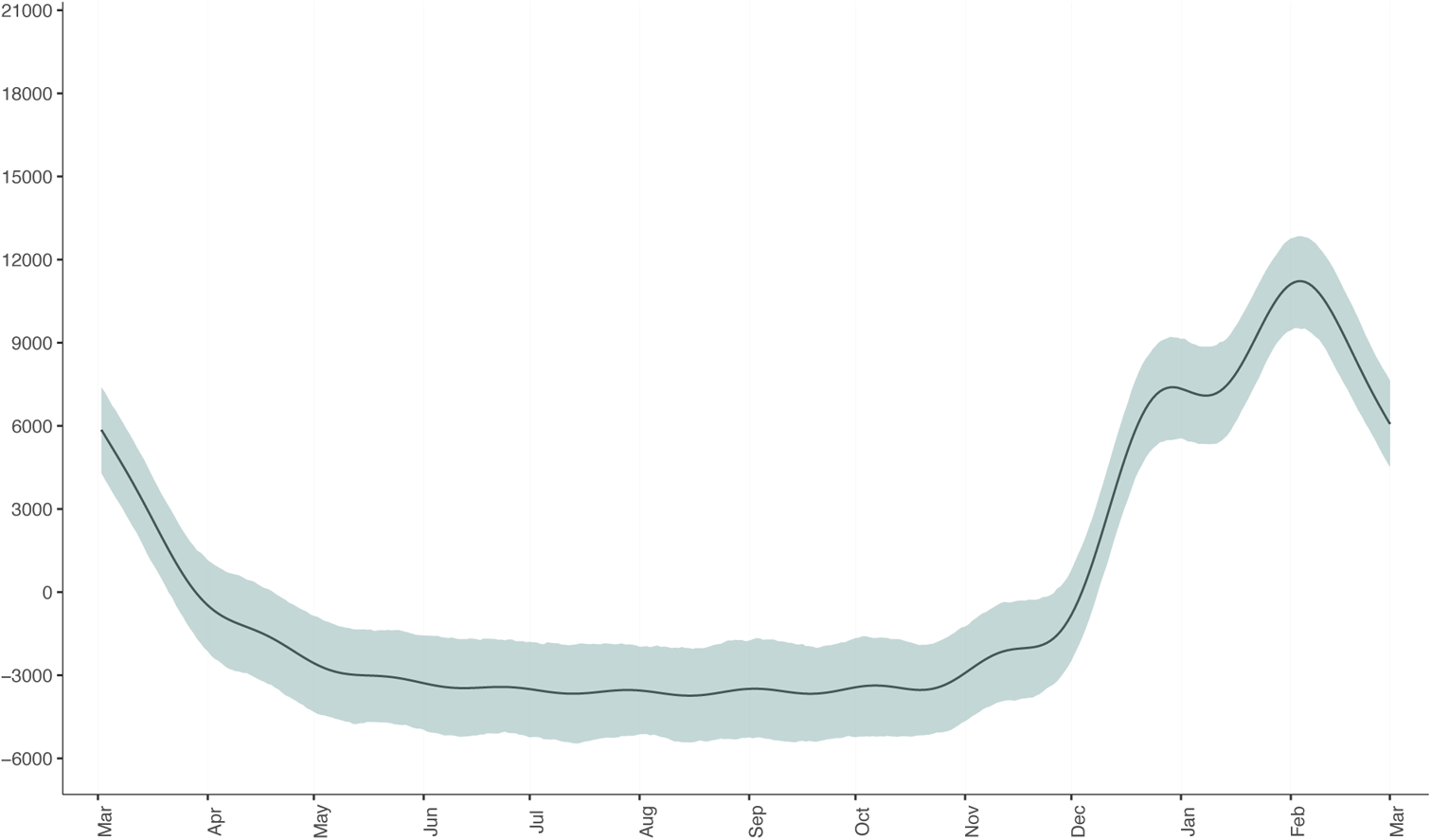
Seasonal component of Influenza cases from the Prophet decomposed model with Markov chain Monte Carlo Simulated 95% uncertainty intervals, Mar 1, 2014 – Apr 10, 2020, holiday-adjusted, United States. Data Source: US CDC

**Figure s15.**
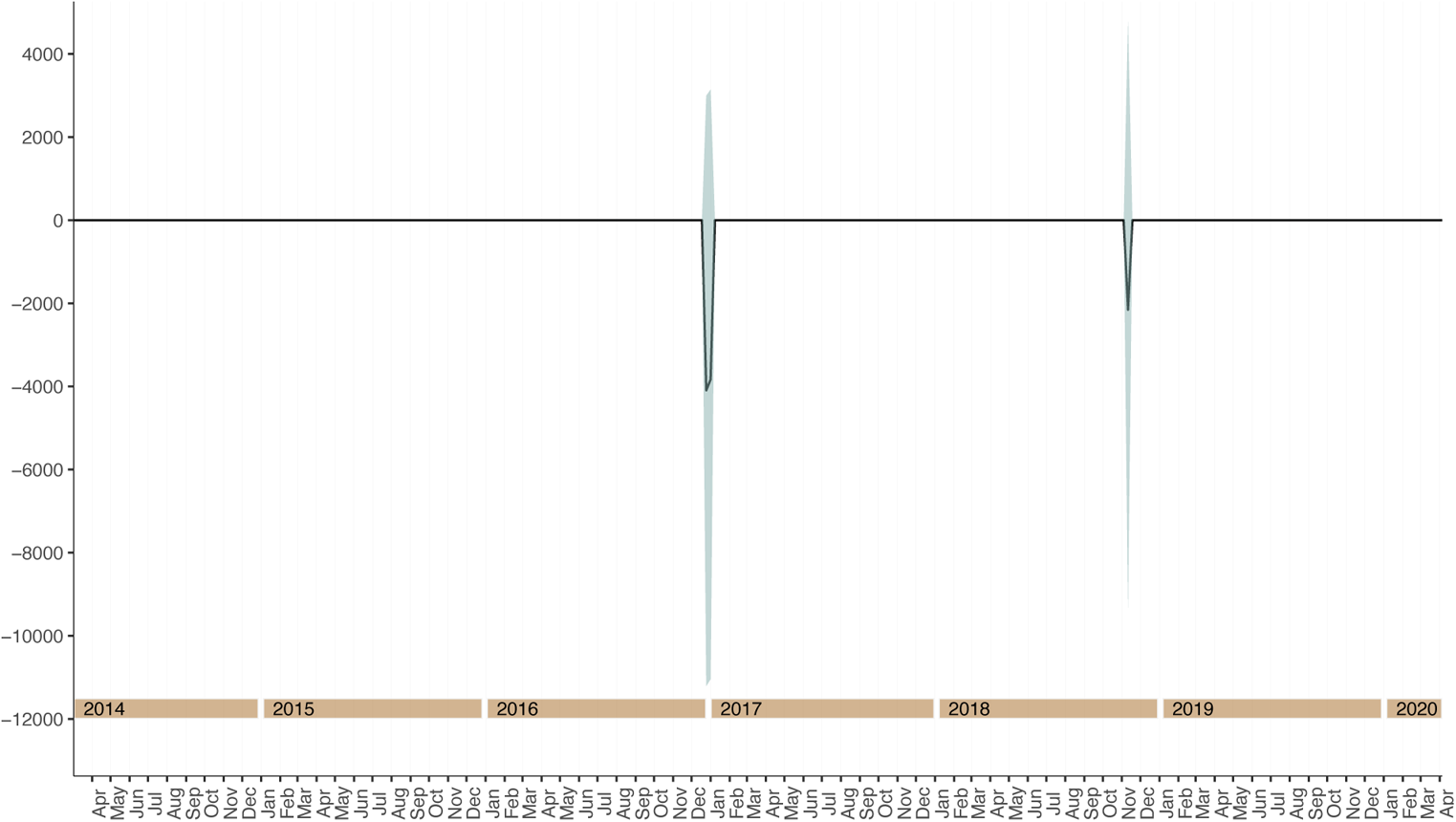
Holiday component of Influenza cases from the Prophet decomposed model with Markov chain Monte Carlo Simulated 95% uncertainty intervals, Mar 1, 2014 – Apr 10, 2020, holiday-adjusted, United States. Data Source: US CDC

**Figure s16.**
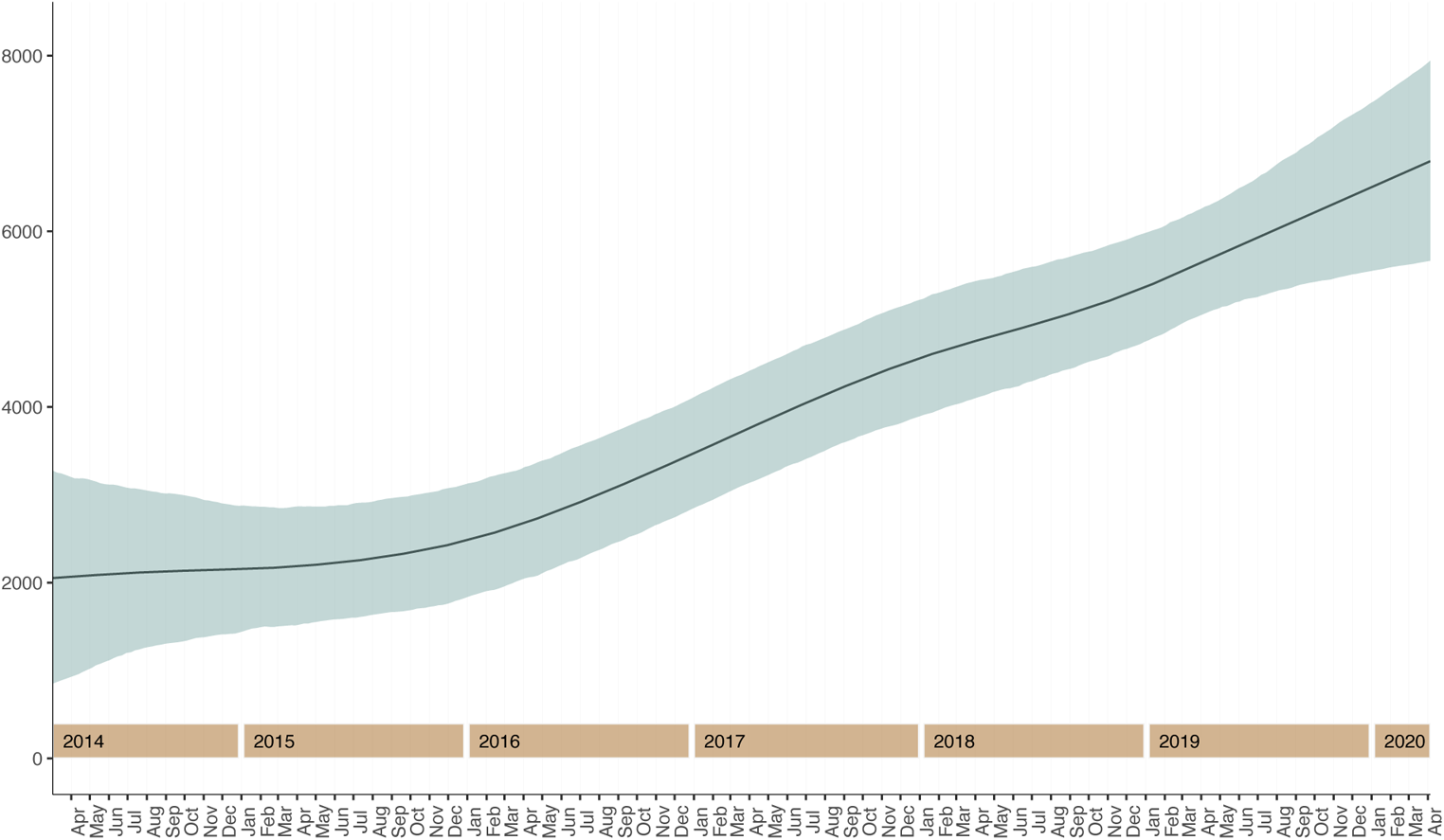
Trend component of Influenza cases from the Prophet decomposed model with Markov chain Monte Carlo Simulated 95% uncertainty intervals, Mar 1, 2014 – Apr 10, 2020, holiday-adjusted, United States. Data Source: US CDC

**Figure s17.**
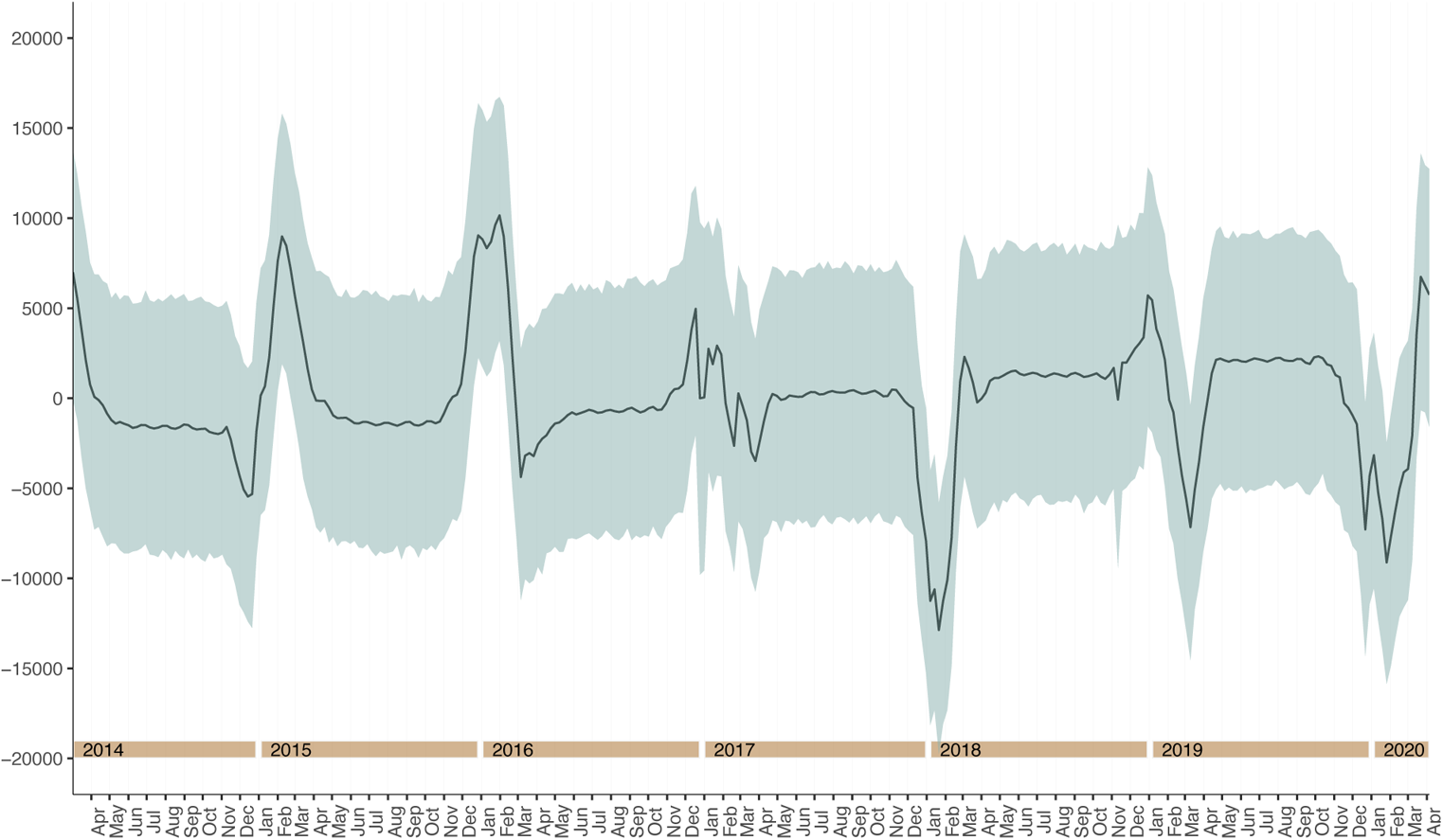
Residual component of Influenza cases from the Prophet decomposed model with Markov chain Monte Carlo Simulated 95% uncertainty intervals, Mar 1, 2014 – Apr 10, 2020, holiday-adjusted, United States. Data Source: US CDC

